# Socioeconomic and geographic correlates of intimate partner violence in Sri Lanka: Analysis of the 2016 Demographic and Health Survey

**DOI:** 10.1101/2021.03.21.21254059

**Authors:** Piumee Bandara, Duleeka Knipe, Sithum Munasinghe, Thilini Rajapakse, Andrew Page

**Affiliations:** Translational Health Research Institute, Western Sydney University, New South Wales, Australia; South Asian Clinical Toxicology Research Collaboration, Faculty of Medicine, University of Peradeniya, Sri Lanka; Population Health Sciences, Bristol Medical School, University of Bristol, Bristol, UK; Department of Psychiatry, Faculty of Medicine, University of Peradeniya, Peradeniya, Sri Lanka

**Keywords:** intimate partner violence, domestic violence, Sri Lanka, socioeconomic position, poverty, post-conflict, multi-level modelling

## Abstract

Intimate partner violence (IPV) is a serious public health issue and violation of human rights. The prevalence of IPV in South Asia is especially pronounced. This is the first study in Sri Lanka to examine the associations between socioeconomic position (SEP), geographical factors and IPV using nationally representative data. Data collected from Sri Lanka’s 2016 Demographic and Health Survey were analysed using multilevel logistic regression techniques. A total 16,390 eligible ever-partnered women aged 15-49 years were included in the analysis. Analyses were also stratified by ethnicity, type of violence, neighbourhood poverty and post-conflict residential status for selected variables. Consistent associations were found for low SEP and IPV. Lower educational attainment among women (OR 2.46 95% CI 1.83-3.30) and their partners (OR 2.87 95% CI 2.06-4.00), financial insecurity (OR 2.17 95% CI 1.92-2.45) and poor household wealth (OR 2.64 95% CI 2.22-3.13) were the socioeconomic factors that showed the strongest association with any IPV, after adjusting for age and religion. These associations predominately related to physical and/or sexual violence, with weak associations for psychological violence. Women living in a post-conflict environment had a higher risk (OR 2.96 95% CI 2.51-3.49) of IPV compared to other areas. Ethnic minority women (Tamil and Moor) were more likely to reside in post-conflict areas and experience poverty more acutely compared to the majority Sinhala women, which may explain the stronger associations for low SEP, post-conflict residence and IPV found among Tamil and Moor women. We found IPV is more likely to occur in the context of socioeconomic disadvantage and post-conflict regions. Further exploratory studies investigating the complex interplay of individual, household and community-level factors occurring in this context is required.

---

Intimate partner violence (IPV) represents a violation of human rights and is a universal public health problem that results in the loss of many lives and many more non-fatal physical and mental health consequences (García-Moreno, Jansen, Ellsberg, Heise, & Watts, 2005). IPV, as defined by the World Health Organization (WHO), refers to any act of violence within an intimate relationship that results in physical, sexual, psychological, or economic harm and suffering (WHO, 2013). According to estimates by the WHO, one in three women worldwide have experienced violence by an intimate partner at some point in their lifetime (WHO, 2013). The burden of lifetime physical and/or sexual violence among ever-partnered women is higher in low and middle-income countries (LMIC), and particularly acute in South Asia where 42% of ever-partnered women have experienced IPV, compared to 23% in high-income countries (WHO, 2013).

Social and economic conditions determine levels of material wealth, psychosocial support, and behavioural choices, often resulting in a social gradient in health, where lower socioeconomic position (SEP) is associated with worse health (CSDH, 2008). There is increasing evidence from LMIC to suggest that IPV is more likely to occur in the context of poverty (Jewkes et al., 2017; Vyas & Watts, 2009). Studies from Asia have reported a 3-fold increased risk of IPV among women with lower socioeconomic backgrounds, measured using a range of socioeconomic indicators (Ackerson & Subramanian, 2008; Dalal & Lindqvist, 2012; Jewkes et al., 2017; Ram et al., 2019).

While Sri Lanka shares similar characteristics to Asian countries, it ranks highest in Asia with respect to tertiary education of women, with women outnumbering men in bachelor degree enrolment (Chapman & Chien, 2014). Sri Lanka also reports strong maternal health indicators, with a maternal mortality ratio of 36 per 100,000 live births, considerably lower than neighbouring India (145/100,000) and Bangladesh (173/100,000) (WHO, 2019). Despite this, a 2015 scoping review estimated that up to 35% of women reported experiencing IPV in their lifetime (Guruge, Jayasuriya-Illesinghe, Gunawardena, & Perera, 2015). As a middle-income country and emerging economy, Sri Lanka is undergoing a period of increasing urbanisation and transition in social and gender norms. Sri Lanka has also experienced a history of violent insurrections and is still recovering from the social, economic and public health impacts of a protracted civil war that was concentrated in the Northern and Eastern provinces, areas largely populated by Tamil and Moor ethnic minority groups. Despite the war ending in 2009, ethnic/religious minority groups, particularly those living in post-conflict areas also continue to disproportionately experience violence and poverty (Mahadevan & Jayasinghe, 2019). Exposure to collective violence and trauma has been previously shown to be associated with an increased likelihood of perpetration of IPV and victimisation in Sri Lanka and other LMIC settings (Fisher, 2010; Fonseka, Minnis, & Gomez, 2015; Fulu et al., 2017). Given these unique socioeconomic and post-conflict conditions, context specific information regarding the relationship between SEP, conflict exposure and IPV is needed to inform prevention and priority-setting.

In 2016, the Sri Lanka Demographic and Health Survey (SLDHS) introduced, for the first time, questions on experience of physical, sexual, and psychological IPV in the previous 12-months among ever-partnered women aged 15-49 years (DCS, 2016). The SLDHS 2016 is the only nationally representative sample of IPV in Sri Lanka and to date, no studies have comprehensively explored the possible association between conflict exposure, SEP and IPV. Using the SLDHS we aimed to examine 1) the association between individual, household, and community-level socioeconomic factors, post-conflict residence and any IPV; 2) the extent to which associations with SEP and post-conflict residence differ by ethnicity; 3) SEP and post-conflict associations with physical/sexual abuse and separately with psychological abuse; 4) whether the association between individual and partner education and IPV is modified by neighbourhood poverty; and 5) if associations between SEP, ethnicity and IPV are modified by post-conflict residence.

## Methods

### Study setting

Sri Lanka is an island nation situated in the Indian ocean. Sri Lanka has a dense population of approximately 20 million, with 77% residing in rural areas, 18% in urban areas and 4% in the plantation sector (DCS, 2012). The majority of Sri Lankans identify as Sinhalese (75%), followed by Tamil (11%) and Moor (9%). Religious adherence is closely linked to ethnicity, with most Sinhalese people identifying as Buddhist, most Tamils as Hindu, and Moors as Muslim. Between 1989-2009, Sri Lanka experienced a civil war between government forces and Tamil separatists, with the conflict largely concentrated in the Northern (94% Tamil population) and Eastern (40% Tamil and 37% Moor population) provinces (DCS, 2012). In addition, the Eastern province was the region worst affected by the 2004 tsunami in terms of human lives lost and households damaged (J. S. Bandara & Naranpanawa, 2007).

### Data

This study used data from the 2016 SLDHS. Detailed data collection methods and questionnaire design are described in the 2016 SLDHS Report (DCS, 2016). In brief, the survey used a two-stage stratified cluster sampling technique to select study participants. At the first stage, enumeration areas (census blocks), were stratified by district and then by sector (urban, rural and estate). Census blocks were then selected with probability proportional to size, described hereafter as the primary sampling units (PSUs). In stage two, a fixed number of households from each PSU were selected by equal probability systematic sampling. Between May to November 2016, only one eligible ever-partnered woman (i.e. married, living with a partner, or separated/divorced/husband died) per household was selected for the module in accordance with the WHO guidelines for the ethical collection of information on DV. The questionnaire was implemented in private and additional consent was obtained, informing the participant of the sensitivity of the questions and that responses would remain confidential (DCS, 2016).

The DHS survey was undertaken by the Sri Lankan Department of Census and Statistics with financial support from the Ministry of Health, Nutrition and Indigenous Medicine in collaboration with the World Bank. Technical assistance was provided by the Inner City Fund International (Inc.), USA.

### Outcome variables

The primary outcome measure was defined as exposure to any type of IPV by husband or partner in the last 12 months preceding the survey (versus no exposure to IPV in the previous year). Binary outcome variables were also created specifically for physical/sexual violence and for psychological violence. Physical violence was defined as being ‘slapped’, ‘pushed’, ‘beaten with an object’, ‘strangled’ or ‘burned’ in the past year. Sexual violence was defined as ‘forced sex’. Due to the limited number of sexual violence cases occurring without any other form of abuse (2% of IPV cases) and the majority of sexual violence cases coinciding with physical violence (74%), a composite physical/sexual violence variable was also created, comparing women who have reported having experienced any act of physical and/or sexual violence with or without psychological violence in the past year versus women who did not report any IPV. Psychological violence was categorised as being prevented from leaving home, or experiencing belittlement or serious offence, without physical and/or sexual violence in the past year.

### Explanatory variables

Demographic factors, including respondent age, marital status, religion, and ethnicity were examined. Factors that have been previously identified as markers of SEP in the literature and associated with IPV were also included in this analysis (Abramsky et al., 2011; Guruge et al., 2015; Jewkes et al., 2017). SEP was defined in this study as the level of material deprivation at the individual, household and community level. At the individual level, SEP measures included educational attainment and occupational status of the woman and partner, and financial insecurity – an income-related measure based on the question ‘do you have enough money for the daily expenses of your house?’ Partner related questions were only administered to women currently married or living with a partner, and financial insecurity question limited to women unemployed. Household wealth was also examined using the wealth index quintile, a composite variable that uses a combination of household ownership of durable goods and housing characteristics (DCS, 2016). A binary community-level poverty variable was also generated using the wealth index quintile to represent neighbourhoods with a high proportion (above the median) of women living in households in the poorest two quintiles versus a low proportion of women living in poorest quintiles.

### Statistical analysis

The data analysis plan was developed and registered *a priori* (P. Bandara, Knipe, & Page, 2020). Characteristics of those who did not respond to the IPV questionnaire were compared with respondents using weighted frequency cross-tabulations, and chi square tests conducted to assess heterogeneity and potential selection bias. The level of missing data was examined for all study variables and statistical evidence of a difference in the level of missing data between women who reported IPV versus no IPV was assessed using a chi square test. Analyses were conducted on all available data. Sensitivity analysis were conducted using information from individuals with complete data to assess robustness of the models.

Weighted column frequency and percentages of IPV by study factors were estimated for overall and stratified analyses. District-level weighted prevalence of IPV using the total number of respondents for each district as the denominator, was calculated and transposed to a choropleth map. IPV prevalence intervals for the map were defined equally into five classes. The middle interval defined in this study is consistent with reports of IPV in Sri Lanka, ranging between 25% to 35% (Guruge et al., 2015). District-level proportion of non-Sinhalese respondents, household poverty (lowest two wealth quintiles), and lower educational attainment (no schooling or primary education) were also transposed to choropleth maps. The administrative boundary layer package was used to generate the map (GADM, 2015).

To account for the hierarchical structure of the data, the association between study factors and IPV were examined using a series of weighted multilevel (two levels) logistic regression models, adjusting for age and religion, previously identified as correlates in this context (Guruge et al., 2015; Jayasuriya, Wijewardena, & Axemo, 2011). Due to collinearity between SEP variables, only minimally adjusted models were presented. Associations specifically for socioeconomic factors and post-conflict residence with any IPV were then stratified by major ethnic groups (Sinhala, Tamil, and Moor), given ethnic minority communities (Moor and Tamil) experience higher rates of poverty and are more likely to reside in post-conflict areas than the Sinhala majority (Mahadevan & Jayasinghe, 2019). Interaction terms were added to the model to determine if potential associations statistically differed by ethnicity. In addition, due to the scarcity of evidence in distinguishing the effects of SEP on different forms of abuse and potentially important differences by abuse type, analyses using the overall sample were then stratified by physical/sexual abuse and psychological abuse.

The proportion of unadjusted total variance in IPV attributable to community-level variance (variance partition coefficient [VPC]) was estimated using the latent variable method described by Merlo et al. (2006). Given the strong association between education and IPV previously shown in Asia (Jewkes et al., 2017), we considered it important to examine the contextual effect of community-level poverty on the association between individual and partner education and IPV. Associations for individual and partner education with IPV were stratified by the level of neighbourhood poverty (high versus low), and cross-level interaction terms added into unweighted models to test for statistical differences. Finally, a supplementary analysis was conducted stratifying the overall sample by post-conflict residence given the effect of exposure to civil conflict on both SEP and IPV (Guruge et al., 2017; Mahadevan & Jayasinghe, 2019). Interaction terms were added to the model to determine if potential associations statistically differed by post-conflict residential status. Women’s occupational status was collapsed into an additional binary (employed vs. unemployed) variable and the association with any IPV was analysed post-hoc to allow for comparisons with existing literature.

All analyses were conducted in Stata (version 15.1, Stata Corp, College Station, TX, USA). Weighted cross-tabulations were estimated using ‘svy’ for counts and percentages. The choropleth map was generated using the ‘spmap’ command. The ‘melogit’ command was used for multilevel regression analyses, and ‘estat icc’ command for intraclass correlation/VPC.

## Results

In total, 18,302 ever-partnered women aged 15-49 years were interviewed, of which 16,390 women (90% response rate) completed the IPV questionnaire. According to the SLDHS 2016 report, 7% were excluded on the basis of privacy issues and 2% declined to participate (DCS, 2016). Less than 1% of participants who chose not to reply to all IPV questions administered were excluded from the analysis. Characteristics of respondents versus non-respondents are summarised in Supplementary Table 1. Compared to respondents, those who did not respond were more likely to be under 35 years, come from a Moor background and identify as Islamic or Christian. A higher proportion of non-respondents (15%) were divorced, separated or their husband had died, compared to respondents (5%; p<0.001). No schooling or primary education of women and her partner, and financial insecurity was higher among those who did not respond compared to respondents. Respondents were more likely to have a partner employed in a professional occupation and reside in a post-conflict region compared to non-respondents. Those who responded from post-conflict areas were more likely to be Tamil or Moor, collectively constituting 83% of the post-conflict region. There was no statistical evidence of differences between respondents and non-respondents for individual occupational status, household wealth, neighbourhood poverty, and residential sector.

Most variables examined had no missing data except for partner education (11%) and partner occupational status (16%). The level of missing data (weighted N=2556, 16%) was higher among those without IPV (16%) than with IPV (14%; p=0.003).

### Prevalence and associations with IPV

Exposure to any IPV in the previous 12 months was reported by 17% (95% CI 16% - 18%) of women overall. At least a third of women in post-conflict areas reported IPV in the past year and the highest IPV prevalence was found in the post-conflict districts of Kilinochchi (50% 95% CI 45% - 56%) and Batticaloa (50% 95% CI 45% - 55%), located in the Northern and Eastern provinces, respectively (Figure 1; Supplementary Table 2). Among those who experienced IPV, the most common abuse (with or without any other form of abuse) was psychological (79% 95% CI 78% - 81%), followed by physical (55% 95% CI 52% - 57%) and sexual abuse (15% 95% CI 14% - 17%), and 10% (95% CI 9% - 11%) experienced all three forms of abuse. Psychological abuse without any physical/sexual violence comprised 42% (95% CI 39% - 44%) of all IPV cases.

**Figure 1.**
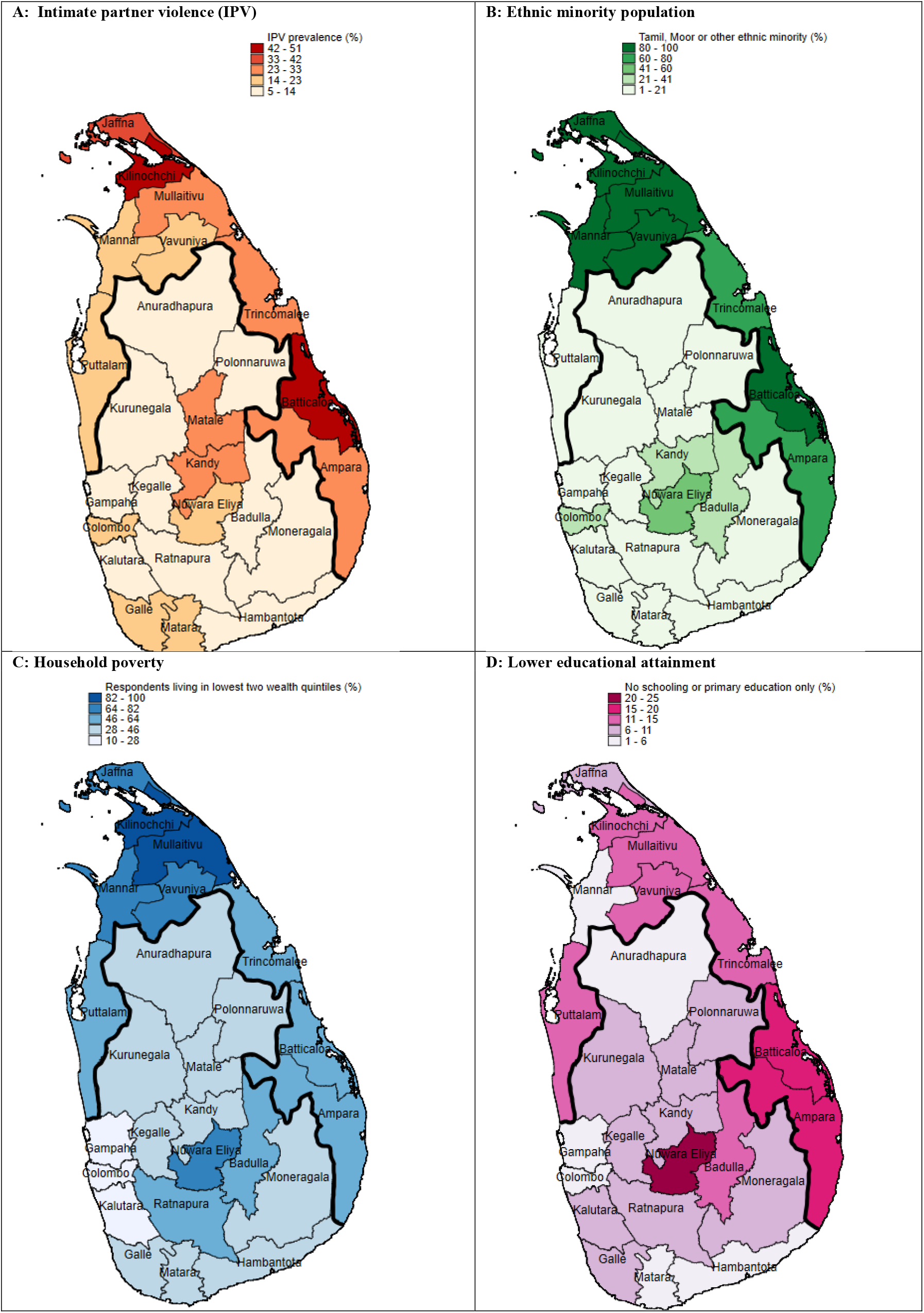
District-level proportion of ever-partnered women aged 15-49 years with: (A) Intimate partner violence (IPV) in the previous 12-months (B) Ethnic minority background (Tamil, Moor, other) (C) Poor household wealth (lowest two quintiles) (D) No schooling or primary educational attainment. Districts above the bold line are post-conflict areas. Data: 2016 Sri Lanka Demographic and Health Survey respondents to the IPV questionnaire (N=16,390).

Women who reported IPV were more likely to be 35 years and older (OR 1.22 95% CI 1.11-1.35) and divorced, separated or widowed (OR 1.60 95% CI 1.31-1.96) than married (Table 1). Women with no schooling or primary education were more likely to experience any IPV (OR 2.5 95% CI 1.83-3.30) than women with a higher education. A similar pattern was found among women whose partner had no schooling or primary education (OR 2.87 95% CI 2.06-4.00), compared to those with a higher education. Women employed in manual labour were also more likely to report any IPV (OR 1.37 95% CI 1.10-1.72) compared to women in professional occupations, and this pattern was similar for women whose partner was employed in manual labour (OR 1.20 95% CI 1.03-1.41) or unemployed (OR 1.47 95% CI 1.05-2.07). Post-hoc analysis of employment vs. unemployment showed a protective effect for unemployment (OR 0.89 95% CI 0.81-0.99). Financial insecurity among unemployed women was similarly associated with a higher likelihood of any IPV (OR 2.17 95% CI 1.92-2.45). Women in the lowest wealth quintile experienced the highest odds (OR 2.39 95% CI 2.03-2.80) compared to those in the highest quintile.

**Table 1.** Intimate partner violence (IPV) in the past year among ever-partnered women aged 15-49 years, Sri Lanka: weighted distribution with all study factors.

|  | IPV (%) | No IPV (%) | OR (95% CI) |
| --- | --- | --- | --- |
| *Individual level factors* |  |  |  |
| **Age** |  |  |  |
| 15-34 | 1102 (39.9) | 5815 (42.7) | 1.00 |
| 35-49 | 1660 (60.1) | 7817 (57.3) | 1.22 (1.11-1.35) |
| **Ethnicity** |  |  |  |
| Sinhala | 1708 (61.9) | 10829 (79.4) | 1.00 |
| Tamil | 767 (27.8) | 1609 (11.8) | 2.96 (2.18-4.03) |
| Moor | 280 (10.1) | 1141 (8.4) | 2.97 (1.73-5.10) |
| Other | 6 (0.2) | 52 (0.4) | 1.07 (0.42-2.75) |
| **Religion** |  |  |  |
| Buddhist | 1600 (57.9) | 10130 (74.3) | 1.00 |
| Hindu | 600 (21.7) | 1255 (9.2) | 2.96 (2.57-3.43) |
| Islam | 296 (10.7) | 1233 (9.0) | 1.45 (1.18-1.77) |
| Christian/Other | 265 (9.6) | 1012 (7.4) | 1.62 (1.35-1.95) |
| **Marital status** |  |  |  |
| Married | 2476 (89.7) | 12502 (91.7) | 1.00 |
| Living with a man | 91 (3.3) | 571 (4.2) | 0.90 (0.68-1.19) |
| Divorced/Separated/Husband died | 195 (7.1) | 557 (4.1) | 1.60 (1.31-1.96) |
| **Educational attainment** |  |  |  |
| Higher education | 99 (3.6) | 681 (5.0) | 1.00 |
| Secondary education | 2268 (82.1) | 12015 (88.1) | 1.33 (1.03-1.72) |
| No education/primary | 395 (14.3) | 935 (6.9) | 2.46 (1.83-3.30) |
| **Partner's educational attainment\*** |  |  |  |
| Higher education | 66 (2.9) | 481 (4.2) | 1.00 |
| Secondary education | 1763 (76.8) | 9940 (86.1) | 1.38 (1.02-1.88) |
| No education/primary | 466 (20.3) | 1117 (9.7) | 2.87 (2.06-4.00) |
| **Occupational status** |  |  |  |
| Professional | 174 (6.3) | 1030 (7.6) | 1.00 |
| Service | 249 (9.0) | 1301 (9.5) | 1.10 (0.86-1.42) |
| Manual labour | 530 (19.2) | 2078 (15.2) | 1.37 (1.10-1.72) |
| Unemployed | 1810 (65.5) | 9222 (67.7) | 1.08 (0.89-1.32) |
| **Partner's occupational status\*** |  |  |  |
| Professional | 350 (15.9) | 2080 (19.1) | 1.00 |
| Service | 519 (23.7) | 3092 (28.4) | 0.97 (0.81-1.16) |
| Manual labour | 1241 (56.6) | 5407 (49.6) | 1.20 (1.03-1.41) |
| Unemployed | 83 (3.8) | 314 (2.9) | 1.47 (1.05-2.07) |
| **Enough income for daily living expenses\*\*** |  |  |  |
| Yes | 1177 (65.1) | 7307 (79.2) | 1.00 |
| No | 632 (34.9) | 1916 (20.8) | 2.17 (1.92-2.45) |
| *Household level factors* |  |  |  |
| **Household wealth index** |  |  |  |
| 1 - Richest | 385 (13.9) | 2829 (20.8) | 1.00 |
| 2 | 443 (16.0) | 2993 (22) | 1.12 (0.94-1.33) |
| 3 | 480 (17.4) | 2959 (21.7) | 1.25 (1.05-1.50) |
| 4 | 595 (21.5) | 2706 (19.8) | 1.67 (1.40-1.99) |
| 5 - Poorest | 859 (31.1) | 2144 (15.7) | 2.64 (2.22-3.13) |
| *Geographical and community level factors* |  |  |  |
| **Sector** |  |  |  |
| Urban | 513 (18.6) | 2049 (15) | 1.00 |
| Rural | 2139 (77.5) | 11064 (81.2) | 0.89 (0.76-1.04) |
| Estate | 110 (4.0) | 518 (3.8) | 0.43 (0.32-0.59) |
| **Province** |  |  |  |
| Western | 513 (18.6) | 3621 (26.6) | 1.00 |
| Central | 490 (17.8) | 1579 (11.6) | 2.22 (1.83-2.70) |
| Southern | 324 (11.7) | 1673 (12.3) | 1.46 (1.19-1.79) |
| Northern | 276 (10) | 516 (3.8) | 2.93 (2.29-3.74) |
| Eastern | 523 (18.9) | 965 (7.1) | 3.85 (3.10-4.78) |
| North-Western | 249 (9) | 1808 (13.3) | 0.99 (0.81-1.21) |
| North-Central | 103 (3.7) | 1139 (8.4) | 0.68 (0.52-0.91) |
| Uva | 100 (3.6) | 980 (7.2) | 0.73 (0.56-0.94) |
| Sabaragamuwa | 183 (6.6) | 1351 (9.9) | 0.98 (0.78-1.23) |
| **Neighbourhood poverty** |  |  |  |
| Low | 1286 (46.6) | 7817 (57.3) | 1.00 |
| High | 1476 (53.4) | 5814 (42.7) | 1.31 (1.16-1.48) |
| **Post-conflict residence** |  |  |  |
| No | 1962 (71.1) | 12150 (89.1) | 1.00 |
| Yes | 799 (28.9) | 1481 (10.9) | 2.96 (2.51-3.49) |

OR = Odds Ratio adjusted for age and religion; CI = Confidence Interval.
\*Women divorced/separated/husband died were not asked this question.
\*\*Women working in a job or business were not asked this question.

At a broader contextual level, 21% of the total variation in IPV (unadjusted) was attributed to differences between communities, indicating the importance of community-level factors in influencing variations in the likelihood of IPV. Compared to urban areas, living in the estate sector was protective against IPV (OR 0.43 95% CI 0.32-0.59). Women residing in neighbourhoods with higher poverty were more likely to report any IPV (OR 1.31 95% CI 1.16-1.48) than women in less impoverished areas, and women residing in post-conflict regions were more likely to report any IPV than those living outside post-conflict regions (OR 2.96 95% CI 2.51-3.49).

Minority ethnic (Tamil and Moor) and religious (Hindu, Christian/other and Muslim) backgrounds – sub-populations that experience higher rates of poverty and exposure to conflict – were more likely to report any IPV compared to Sinhala women (Table 1).

Stratified analysis by ethnicity showed evidence of statistical differences for associations with post-conflict residence (p<0.001) and most socioeconomic indicators, except household wealth and financial insecurity (Table 2). Point estimates were larger for Moor women whose partner had no schooling or primary education (OR 11.39 95% CI 3.47-37.35), compared to Sinhala (OR 2.97 95% CI 1.95-4.52) and Tamil women (OR 2.04 95% CI 1.11-3.19; interaction p value=0.003) (Table 2).

**Table 2.** Intimate partner violence (IPV) in the past year among ever-partnered women aged 15-49 years, Sri Lanka: weighted prevalence and adjusted associations with socioeconomic factors and post-conflict residence, stratified by major ethnic groups.

|  | Sinhala |  |  | Tamil |  |  | Moor |  |  | P value interaction |
| --- | --- | --- | --- | --- | --- | --- | --- | --- | --- | --- |
|  | IPV (%) | No IPV (%) | OR (95% CI) | IPV (%) | No IPV (%) | OR (95% CI) | IPV (%) | No IPV (%) | OR (95% CI) |  |
| *Individual & partner factors* |  |  |  |  |  |  |  |  |  |  |
| **Educational attainment** |  |  |  |  |  |  |  |  |  |  |
| Higher education | 68 (4.0) | 563 (5.2) | 1.00 | 24 (3.1) | 71 (4.4) | 1.00 | 4 (1.6) | 44 (3.8) | 1.00 | <0.001 |
| Secondary education | 1473 (86.2) | 9765 (90.2) | 1.26 (0.93-1.70) | 570 (74.4) | 1208 (75.1) | 1.51 (0.89-2.54) | 220 (78.5) | 995 (87.2) | 2.03 (0.76-5.47) |  |
| No schooling/primary education | 167 (9.8) | 501 (4.6) | 2.79 (1.93-3.99) | 172 (22.5) | 330 (20.5) | 1.85 (1.04-3.29) | 56 (19.9) | 103 (9.0) | 4.80 (1.74-13.24) |  |
| **Partner's educational attainment** |  |  |  |  |  |  |  |  |  |  |
| Higher education | 41 (2.8) | 373 (4.0) | 1.00 | 21 (3.4) | 58 (4.6) | 1.00 | 3 (1.2) | 47 (5.1) | 1.00 | 0.003 |
| Secondary education | 1160 (80.9) | 8194 (88.1) | 1.30 (0.89-1.92) | 427 (68.4) | 937 (73.5) | 1.37 (0.77-2.43) | 171 (74.6) | 775 (84.3) | 4.05 (1.31-12.53) |  |
| No schooling/primary education | 234 (16.3) | 737 (7.9) | 2.97 (1.95-4.52) | 176 (28.2) | 280 (21.9) | 2.04 (1.11-3.19) | 55 (24.1) | 97 (10.6) | 11.39 (3.47-37.35) |  |
| **Occupational status** |  |  |  |  |  |  |  |  |  |  |
| Professional | 145 (8.5) | 906 (8.4) | 1.00 | 25 (3.3) | 77 (4.8) | 1.00 | 3 (1.0) | 39 (3.4) | 1.00 | <0.001 |
| Service | 177 (10.4) | 1091 (10.1) | 1.06 (0.80-1.40) | 55 (7.1) | 157 (9.7) | 1.10 (0.65-1.87) | 17 (5.9) | 45 (3.9) | 4.82 (1.16-19.91) |  |
| Manual labour | 348 (20.4) | 1639 (15.1) | 1.33 (1.03-1.70) | 141 (18.4) | 348 (21.6) | 1.33 (0.80-2.20) | 40 (14.3) | 85 (7.4) | 6.08 (1.63-22.70) |  |
| Unemployed | 1038 (60.8) | 7192 (66.4) | 0.94 (0.76-1.17) | 546 (71.2) | 1027 (63.8) | 1.64 (1.03-2.62) | 221 (78.8) | 972 (85.2) | 3.18 (0.88-11.48) |  |
| **Partner's occupational status\*** |  |  |  |  |  |  |  |  |  |  |
| Professional | 251 (18.7) | 1767 (20.3) | 1.00 | 89 (14.3) | 156 (12.4) | 1.00 | 8 (3.6) | 147 (16.2) | 1.00 | <0.001 |
| Service | 360 (26.8) | 2494 (28.7) | 1.00 (0.81-1.23) | 98 (15.7) | 285 (22.6) | 0.64 (0.43-0.96) | 59 (26.6) | 300 (33.0) | 3.84 (1.49-9.92) |  |
| Manual labour | 670 (49.9) | 4173 (48) | 1.11 (0.92-1.34) | 421 (67.7) | 778 (61.6) | 0.95 (0.69-1.33) | 148 (66.9) | 442 (48.7) | 6.30 (2.66-14.87) |  |
| Unemployed | 62 (4.6) | 251 (2.9) | 1.62 (1.10-2.39) | 14 (2.3) | 43 (3.4) | 0.60 (0.30-1.23) | 6 (2.8) | 20 (2.2) | 7.96 (1.94-32.71) |  |
| **Enough income for daily living expenses\*\*** |  |  |  |  |  |  |  |  |  |  |
| Yes | 641 (61.7) | 5692 (79.1) | 1.00 | 376 (68.8) | 819 (79.7) | 1.00 | 158 (71.6) | 769 (79.1) | 1.00 | 0.13 |
| No | 397 (38.3) | 1501 (20.9) | 2.37 (2.03-2.77) | 170 (31.2) | 208 (20.3) | 2.00 (1.61-2.51) | 63 (28.4) | 203 (20.9) | 1.79 (1.22-2.64) |  |
| *Household level factors* |  |  |  |  |  |  |  |  |  |  |
| **Household wealth index** |  |  |  |  |  |  |  |  |  |  |
| 1 - Richest | 319 (18.7) | 2405 (22.2) | 1.00 | 29 (3.8) | 139 (8.6) | 1.00 | 34 (12.1) | 259 (22.7) | 1.00 | 0.11 |
| 2 | 328 (19.2) | 2518 (23.3) | 1.02 (0.84-1.24) | 61 (8.0) | 181 (11.2) | 1.64 (0.99-2.71) | 54 (19.1) | 286 (25.0) | 1.51 (0.84-2.70) |  |
| 3 | 344 (20.1) | 2526 (23.3) | 1.09 (0.9-1.33) | 74 (9.6) | 195 (12.1) | 1.76 (1.07-2.92) | 62 (22.1) | 234 (20.5) | 2.34 (1.31-4.16) |  |
| 4 | 366 (21.4) | 2145 (19.8) | 1.42 (1.16-1.74) | 164 (21.4) | 336 (20.9) | 2.50 (1.59-3.94) | 63 (22.5) | 219 (19.2) | 2.40 (1.38-4.17) |  |
| 5 - Poorest | 352 (20.6) | 1235 (11.4) | 2.41 (1.97-2.96) | 439 (57.2) | 758 (47.1) | 3.20 (2.08-4.94) | 68 (24.2) | 143 (12.5) | 4.07 (2.39-6.92) |  |
| *Community level factors* |  |  |  |  |  |  |  |  |  |  |
| **Neighbourhood poverty** |  |  |  |  |  |  |  |  |  |  |
| Low | 1025 (60.0) | 6692 (61.8) | 1.00 | 122 (15.9) | 366 (22.7) | 1.00 | 134 (47.7) | 720 (63.1) | 1.00 | <0.001 |
| High | 683 (40.0) | 4137 (38.2) | 1.09 (0.95-1.26) | 645 (84.1) | 1243 (77.3) | 1.59 (1.17-2.15) | 147 (52.3) | 421 (36.9) | 2.31 (1.48-3.60) |  |
| **Post-conflict residence** |  |  |  |  |  |  |  |  |  |  |
| No | 1655 (96.9) | 10497 (96.9) | 1.00 | 194 (25.2) | 765 (47.5) | 1.00 | 110 (39.4) | 840 (73.6) | 1.00 | <0.001 |
| Yes | 54 (3.1) | 332 (3.1) | 1.07 (0.80-1.43) | 573 (74.8) | 844 (52.5) | 3.03 (2.38-3.87) | 170 (60.6) | 301 (26.4) | 4.97 (3.33-7.41) |  |

OR = Odds Ratio adjusted for age and religion; CI = Confidence Interval. P value for interaction relates to differences by ethnicity.
\*Women divorced/separated/husband died were not asked this question.
\*\*Women working in a job or business were not asked this question.

Stratification by type of abuse outcome showed that associations with most measures of low SEP related to physical/sexual violence but not psychological violence. However, women employed in a professional occupation were more at risk of psychological abuse than women in lower wage occupations(Table 3). No marked differences in the magnitude of the association for post-conflict residence were found between psychological and physical/sexual violence (Table 3). Stratification by neighbourhood poverty level showed statistical evidence that the association between IPV and individual education were modified by neighbourhood poverty (p<0.001) but not for partner education (p=0.23) (Table 4). Finally, analyses stratified by post-conflict residence showed statistical evidence that associations for women’s and partner’s occupational status, and neighbourhood poverty with IPV varied by post-conflict residence (Supplementary Table 2). Point estimates for manual labour occupations (OR 2.14 95% CI 1.24-3.70) and unemployment (OR 1.95 95% CI 1.17-3.27) were magnified among women living in post-conflict areas, compared to women living in other regions. The inverse was found for partner occupation, with estimates attenuated for post-conflict regions compared to other regions.

**Table 3.** Physical/sexual violence and psychological violence in the past year among ever-partnered women aged 15-49 years, Sri Lanka: adjusted associations with socioeconomic factors and post-conflict residence.

|  | Physical/sexual violence |  |  | Psychological violence |  |  |
| --- | --- | --- | --- | --- | --- | --- |
|  | Yes<br>N (%) | No<br>N (%) | OR (95% CI) | Yes<br>N (%) | No<br>N (%) | OR (95% CI) |
| **Educational attainment** |  |  |  |  |  |  |
| Higher education | 30 (1.8) | 681 (5.0) | 1.00 | 69 (6) | 681 (5) | 1.00 |
| Secondary education | 1299 (80.4) | 12015 (88.1) | 2.47 (1.63-3.75) | 969 (84.6) | 12015 (88.1) | 0.83 (0.61-1.13) |
| No schooling/primary education | 287 (17.8) | 935 (6.9) | 5.42 (3.48-8.45) | 108 (9.4) | 935 (6.9) | 1.06 (0.62-1.76) |
| **Partner's educational attainment** |  |  |  |  |  |  |
| Higher education | 20 (1.6) | 481 (4.2) | 1.00 | 46 (4.6) | 481 (4.2) | 1.00 |
| Secondary education | 961 (73.2) | 9940 (86.1) | 2.39 (1.46-3.92) | 802 (81.6) | 9940 (86.1) | 0.92 (0.63-1.34) |
| No schooling/primary education | 331 (25.2) | 1117 (9.7) | 6.37 (3.81-10.64) | 135 (13.7) | 1117 (9.7) | 1.20 (0.78-1.85) |
| **Occupational status** |  |  |  |  |  |  |
| Professional | 65 (4.0) | 1030 (7.6) | 1.00 | 109 (9.5) | 1030 (7.6) | 1.00 |
| Service | 143 (8.9) | 1301 (9.5) | 1.71 (1.21-2.43) | 105 (9.2) | 1301 (9.5) | 0.72 (0.52-1.00) |
| Manual labour | 341 (21.1) | 2078 (15.2) | 2.36 (1.72-3.24) | 189 (16.5) | 2078 (15.2) | 0.76 (0.57-1.02) |
| Unemployed | 1067 (66.1) | 9222 (67.7) | 1.70 (1.26-2.27) | 742 (64.8) | 9222 (67.7) | 0.72 (0.56-0.92) |
| **Partner's occupational status\*** |  |  |  |  |  |  |
| Professional | 160 (12.7) | 2080 (19.1) | 1.00 | 190 (20.3) | 2080 (19.1) | 1.00 |
| Service | 257 (20.4) | 3092 (28.4) | 1.00 (0.80-1.28) | 262 (28.1) | 3092 (28.4) | 0.92 (0.72-1.18) |
| Manual labour | 792 (63.0) | 5407 (49.6) | 1.66 (1.36-2.03) | 449 (48) | 5407 (49.6) | 0.80 (0.64-1.00) |
| Unemployed | 49 (3.9) | 314 (2.9) | 1.94 (1.27-2.96) | 33 (3.6) | 314 (2.9) | 1.03 (0.64-1.65) |
| **Enough income for daily living expenses\*\*** |  |  |  |  |  |  |
| Yes | 649 (60.9) | 7307 (79.2) | 1.00 | 932 (81.3) | 11716 (85.9) | 1.00 |
| No | 418 (39.1) | 1916 (20.8) | 2.69 (2.31-3.13) | 215 (18.7) | 1916 (14.1) | 1.76 (1.43-2.17) |
| **Household wealth index** |  |  |  |  |  |  |
| 1 - Richest | 155 (9.6) | 2829 (20.8) | 1.00 | 230 (20.1) | 2829 (20.8) | 1.00 |
| 2 | 220 (13.6) | 2993 (22.0) | 1.38 (1.08-1.77) | 223 (19.5) | 2993 (22) | 0.92 (0.74-1.15) |
| 3 | 268 (16.6) | 2959 (21.7) | 1.71 (1.35-2.18) | 212 (18.5) | 2959 (21.7) | 0.94 (0.74-1.19) |
| 4 | 375 (23.2) | 2706 (19.8) | 2.59 (2.07-3.25) | 220 (19.2) | 2706 (19.8) | 1.00 (0.78-1.28) |
| 5 - Poorest | 598 (37) | 2144 (15.7) | 4.49 (3.60-5.60) | 261 (22.8) | 2144 (15.7) | 1.29 (1.00-1.65) |
| **Community-level poverty** |  |  |  |  |  |  |
| Low | 675 (41.8) | 7817 (57.3) | 1.00 | 611 (53.3) | 7817 (57.3) | 1.00 |
| High | 941 (58.2) | 5814 (42.7) | 1.58 (1.37-1.82) | 535 (46.7) | 5814 (42.7) | 1.00 (0.83-1.21) |
| **Post-conflict residence** |  |  |  |  |  |  |
| No | 1115 (69.0) | 12150 (89.1) | 1.00 | 848 (70.1) | 12150 (89.1) | 1.00 |
| Yes | 501 (31.0) | 1481 (10.9) | 2.90 (2.41-3.50) | 299 (29.9) | 1481 (10.9) | 3.01 (2.37-3.83) |

OR=Odds ratio adjusted for age and religion; CI=Confidence Interval.
\*Women divorced/separated/husband died were not asked this question.
\*\*Women working in a job or business were not asked this question.

**Table 4.** Any intimate-partner violence (IPV) in the past year among ever-partnered women aged 15-49 years, Sri Lanka: adjusted associations with individual and partner education, stratified by neighbourhood poverty.

|  | Poorer neighbourhood |  |  | Wealthier neighbourhood |  |  | Interaction<br>P value |
| --- | --- | --- | --- | --- | --- | --- | --- |
|  | IPV<br>N (%) | No IPV<br>N (%) | OR (95% CI) | IPV<br>N (%) | No IPV<br>N (%) | OR (95% CI) |  |
| **Educational attainment** |  |  |  |  |  |  |  |
| Higher education | 21 (1.4) | 146 (2.5) | 1.00 | 78 (6.1) | 535 (6.9) | 1.00 | <0.001 |
| Secondary education | 1166 (79.0) | 4996 (85.9) | 1.90 (1.17-3.07) | 1103 (85.7) | 7020 (89.8) | 1.13 (0.84-1.52) |  |
| No schooling/primary education | 290 (19.6) | 673 (11.6) | 2.88 (1.71-4.84) | 105 (8.2) | 262 (3.4) | 2.87 (1.93-4.25) |  |
| **Partner's educational attainment** |  |  |  |  |  |  |  |
| Higher education | 19 (1.5) | 76 (1.6) | 1.00 | 47 (4.4) | 405 (6.0) | 1.00 | 0.23 |
| Secondary education | 856 (70.3) | 3982 (82.5) | 1.07 (0.62-1.87) | 907 (84.2) | 5958 (88.7) | 1.40 (0.97-2.02) |  |
| No schooling/primary education | 343 (28.2) | 766 (15.9) | 2.05 (1.15-3.65) | 123 (11.4) | 351 (5.2) | 3.22 (2.08-4.99) |  |

OR=Odds ratio adjusted for age and religion; CI=Confidence Interval.

Sensitivity analysis of associations between all study variables and any IPV using complete case data were consistent with the findings from the main analysis (Supplementary Table 4). Associations for lower education, unemployment and lower wage occupations with IPV were magnified among Moor women, compared to the main analysis. Measures of association for wealth, neighbourhood poverty, and post-conflict residence with psychological abuse were slightly magnified in comparison to the main analysis.

## Discussion

Socioeconomic disadvantage and living in a post-conflict area was associated with past-year IPV among ever-partnered women aged 15-49 years in Sri Lanka. Low SEP associations predominantly related to physical/sexual violence, with weak associations evident among women reporting psychological violence alone, except for post-conflict residence. Women with a lower education were more at risk if they lived in an area of high poverty. Stronger associations for women unemployed or working in manual labour with IPV were found among women living in post-conflict regions, compared to other regions. Moor women were more likely to experience both acute poverty and reside in post-conflict areas than the majority Sinhala population which may explain the magnified associations in this sub-population.

The national prevalence of any past-year IPV reported in this study (17% 95% CI 16% - 18%) is below national 12-month IPV estimates from neighbouring India (26% 95% CI 26%-27%) and Pakistan (25% 95% CI 23% - 26%) (International Institute for Population Sciences, 2017; National Institute of Population Studies & ICF, 2019). In Sri Lanka, unlike its South Asian neighbours, women outnumber men in higher educational attainment, and Sri Lanka ranks highest in the region on gender equality outcomes, with a gender inequality ranking of 86 worldwide, compared to 122 in India, 126 in Bangladesh, and 129 in Pakistan (UNDP, 2019). Advances in women’s empowerment and gender equality may partly explain the comparatively lower estimates in Sri Lanka compared to the wider region. Despite this, strong associations between various indicators of low SEP and IPV were present in this study and is supported by evidence from South Asia and other LMIC (Abramsky et al., 2011; Ackerson, Kawachi, Barbeau, & Subramanian, 2008; Ackerson & Subramanian, 2008; Jewkes et al., 2017; Vyas & Watts, 2009).

### Educational attainment and IPV

Women’s lower education in particular, and men’s education have been found to be associated with a higher risk of IPV, with risk estimates ranging between OR 1.7-2.1 and OR 1.8-3.9 for women’s primary education and men’s primary education respectively (Ackerson & Subramanian, 2008; Ali, Ali, Khuwaja, & Nanji, 2014; Vyas & Watts, 2009).

The protective effect of higher education has been attributed to a range of factors including delayed onset of marriage, an increase in the capability to effectively negotiate conflict and regulate emotions, and increased wealth via enhanced employment opportunities and subsequently a reduced risk of economic stress, a key source of interpersonal conflict (Jewkes et al., 2017; Vyas & Watts, 2009). While higher education is important, there is increasing evidence that integrating teachings around mutual respect and interpersonal skills in school curricula potentially through more comprehensive sexual education may be important strategies to prevent violence (Rollston et al., 2020). Although Sri Lanka has made steps in broadening the curriculum and incorporating life skills education into the syllabus (Clarke, 2010), adolescents continue to report limited sexual and reproductive health knowledge and teachers have reported a disproportionate focus on physiology in the syllabus (Jayasooriya & Mathangasinghe, 2019; Rajapaksa-Hewageegana, Piercy, Salway, & Samarage, 2015). The delivery of more comprehensive sexual education that addresses harmful gender norms in addition to sexual and reproductive health may help equip young people with the social and emotional skills to develop healthy, respectful relationships (Antle, Karam, Christensen, Barbee, & Sar, 2011; Rollston et al., 2020).

### Individual and partner employment and IPV

A number of studies from LMIC have shown IPV is less likely to occur among unemployed women compared to employed women (Abramsky et al., 2011; Bhalotra, Kambhampati, Rawlings, & Siddique, 2020; Jewkes et al., 2017; Naved & Persson, 2005; Terrazas-Carrillo & McWhirter, 2015). This is supported by the present study which showed a protective effect for unemployment compared to employment. Empirical evidence distinguishing the effects of different types of occupations on IPV is limited. However, employment in manual labour has been linked with higher rates of IPV compared to professional occupations in India (Paul, 2016), consistent with results from the present study. For women employed in manual labour occupations, there is likely a complex interplay of sociocultural and socioeconomic factors including lower educational attainment, household economic stress, and partner lower wages.

Moreover, in a country undergoing a transition of social and gender norms, women’s employment has complex implications for IPV. Studies from LMIC suggest women’s increased financial independence may undermine culturally defined gender roles, and violence may be used as an additional means to maintain dominance, particularly in the context of poverty (Ackerson & Subramanian, 2008; Jewkes et al., 2017). Despite this, women’s economic empowerment has important benefits in terms of wider economic growth, gender equality, individual autonomy, and wellbeing of offspring. Notably, in the present study, women experiencing financial insecurity were twice as likely to report IPV. While financial insecurity may be influenced by other factors outside of unemployment such as having a partner employed in a lower wage occupation or husbands/partners deliberately withholding their earnings as a means of control, women who are financially dependent on their partner may find it more difficult to leave an abusive relationship. This is particularly relevant in South Asian settings where access to divorce or separation is limited due to both cultural and legal factors (Bhalotra et al., 2020).

Women whose partners were either unemployed or employed in manual labour, were also at a higher risk of IPV compared to women whose partners were employed in a professional occupation. Lower partner education, wages or unemployment, are by extension associated with poor household wealth in Sri Lanka, as families are predominantly reliant on male income. In the present study, a strong wealth gradient was found for IPV, where poorer household wealth corresponded to an increase in IPV risk. Social scientists have argued that economic stress, due to a lack of material resources including insufficient food, together with potentially reduced capability to regulate emotions (due to lower education) may increase the likelihood of psychological stress and interpersonal conflict (Gibbs et al., 2020). This stress may be compounded among men who feel they cannot fulfil social expectations as the economic provider, which may in turn increase the likelihood of proximal risk factors for the perpetration of violence such as alcohol and substance misuse (Jewkes et al., 2017).

### Neighbourhood poverty, educational attainment and IPV

At the community level, women living in areas of high poverty were at a greater risk of IPV which is consistent with previous studies in LMIC settings (Ackerson & Subramanian, 2008; Naved & Persson, 2005; Vyas & Heise, 2016). Higher rates of poverty are found in the estate sector, largely populated by Indian Tamils, a marginalised ethnic group that disproportionately experience worse health indicators (Jayawardena, 2014). Contrary to local evidence (Infanti et al., 2015; Muzrif, Perera, Wijewardena, Schei, & Swahnberg, 2018), a protective effect was found for women living in the estate sector compared to urban areas. However, urban areas were more likely to be classified as post-conflict which may explain this effect. When compared to the capital Western province, women living in the Central province (which includes major estate populations) were twice as likely to experience IPV. Alcohol and substance misuse occurs across the social gradient, however, areas of socioeconomic disadvantage may facilitate more collective and individual alcohol consumption which in turn may increase IPV risk (Fulu, Jewkes, Roselli, & Garcia-Moreno, 2013; Jayasuriya et al., 2011). Furthermore, higher rates of community violence and IPV have been reported in areas of socioeconomic disadvantage and thus are more likely be accepted as a social norm (VanderEnde, Yount, Dynes, & Sibley, 2012).

Notably, women with a secondary education, relative to higher educated women, were more likely to report IPV if they lived in an area of high poverty compared to a wealthier area. Women in poorer areas with a secondary education were more likely to have a partner with no schooling or primary education and work in manual labour compared to women with a secondary education in wealthier areas (data not shown). The interplay of education differentials, poverty and the transgression of social norms through paid work may explain the higher probability of IPV among these women.

Programs and policies designed to relieve poverty and increase economic empowerment of women should adopt a broader approach to support not only financial security but also the integration of education, for women and their partners, on interpersonal skills, and to challenge the perception and value of women as merely homemakers and men as economic providers. Community-based strategies addressing harmful gender norms in Sri Lanka have shown promise (Herath, Guruge, Fernando, Jayarathna, & Senarathna, 2018), and findings from a cluster randomised trial in Uganda showed a reduction in IPV after community mobilisation (Abramsky et al., 2016).

### Post-conflict residence and IPV

Poverty is highest in the post-conflict region of Sri Lanka largely due to prolonged exposure to war in addition to the 2004 tsunami, events which have collectively resulted in significant deaths, disability, displacement, and loss of livelihood (J. S. Bandara & Naranpanawa, 2007; Mahadevan & Jayasinghe, 2019). The strong association with IPV found in post-conflict areas in the present study is supported by reports conducted in the region and studies conducted in post-war and post-natural disaster settings (Catani, Schauer, & Neuner, 2008; Fisher, 2010; Guruge et al., 2017; Kottegoda, Samuel, & Emmanuel, 2008). It has been argued that prolonged exposure to collective violence may contribute to the normalisation of violence as an acceptable method of conflict resolution (Guruge et al., 2017; Jewkes et al., 2017). Moreover, the impact of war on men’s capability to work and traditional livelihoods (e.g. fishing) has resulted in women becoming more economically active (Guruge et al., 2017). The present study found that women working in lower wage occupations compared to professional occupations had a higher risk of IPV if they lived in a post-conflict region. As previously discussed, in the context of changing gender roles, men may use violence to reassert power. These results highlight the importance of prioritising post-conflict provinces in IPV prevention efforts, and developing tailored trauma informed approaches in this region.

### Effects of SEP and post-conflict residence on physical/sexual abuse versus psychological abuse

Associations between low SEP and IPV predominantly related to physical/sexual violence. Notably, women employed in professional occupations, compared to lower wage occupations were more at risk of psychological violence than physical/sexual violence. In Sri Lanka, conservative attitudes towards gender appear to transgress social boundaries. In a study conducted with 476 medical students, the majority of whom identified as middle-class or upper middle class, 63% agreed that women bear a proportionately larger responsibility for the violence perpetrated against them and a third of students agreed that “wife beating” was justified (Haj-Yahia & de Zoysa, 2007). The persistence of these attitudes across social strata suggest that while higher education and professional employment may protect women from physical violence, this effect does not extend to more covert, non-physical forms of violence such as coercive and controlling behaviours, humiliation and belittlement. Conversely, it is also possible that women with a low SEP may be less likely to recognise more covert forms of abuse as IPV compared to higher educated, wealthier women. Despite this, psychological violence (with or without any other form of violence) had the highest prevalence among all forms of violence reported and has been linked to a range of adverse health consequences, including suicidal behaviours in Sri Lanka and other LMIC settings, (P. Bandara, Page, et al., 2020; Devries et al., 2013; Richardson, Nandi, Jaswal, & Harper, 2020). Further investigation of risk factors for psychological violence is urgently needed to inform prevention.

### Strengths and limitations

This is the first study in Sri Lanka to comprehensively examine the associations between SEP, post-conflict residence and IPV using a nationally representative population-based dataset that provides a sufficient sample size and statistical power. All analyses adhered to a pre-registered protocol. Weighted multilevel analyses were conducted to account for the cluster survey design. A sensitivity analysis using complete case data was also included to assess robustness of the models. An additional strength of this study is that it applied a standard questionnaire which may have minimised the effect of measurement bias. Furthermore, psychological abuse, a neglected area of research in LMIC, was also investigated.

Nevertheless, there are limitations to this study that should be considered. Firstly, the cross-sectional nature of the data does not allow clear temporal relationships between selected exposures (SEP measures) and IPV to be established. The questionnaire was also limited to ever-married women and women living with a partner, potentially excluding a number of unmarried women who have been or are currently in an abusive relationship. Men were also excluded from the IPV questionnaire and the definition of IPV was limited to violence perpetrated by intimate partner or spouse despite reports of strong psychological effects of family violence in this context (Haj-Yahia, Tishby, & de Zoysa, 2009). Future studies should consider broadening the inclusion criteria to include unmarried women (not living with a partner), and explore violence among men as well as the predictors and differential effects of violence perpetrated by other household members versus intimate partners. A sensitivity analysis excluding participants with incomplete data was conducted which showed results were largely consistent with main findings. However, estimates were magnified for Moor women compared to the main analysis, which may indicate differences in the willingness to disclose information relating to SEP variables or IPV among this ethnic group.

A higher proportion of women who were previously married or partnered were found among non-respondents of the IPV questionnaire than respondents. This may have biased the estimates towards the null as the reason for separation may have been IPV. Underreporting may have also occurred due to recall bias (12-month recall of IPV) and stigma surrounding IPV. Sensitivity to public opinion among women of higher social standing may have introduced social desirability bias as these respondents may be more reticent in disclosing abuse. Given that a higher proportion of respondents had a partner employed in a professional occupation and were less likely to have no schooling or primary education compared to non-respondents, it is possible that the strength of the associations with socioeconomic factors may be overestimated. A higher number of respondents were also from post-conflict regions compared to non-respondents. As previously discussed, exposure to civil war has been linked to higher rates of IPV and is also associated with higher rates of poverty, potentially biasing estimates away from the null. Notably, 60% of Tamil and 33% of Moor respondents to the IPV questionnaire lived in a post-conflict region compared to 3% of Sinhala respondents.

Moor women were also less likely to respond to the IPV questionnaire, and those that did respond were more likely to live in poorer households compared to Sinhala respondents. Therefore, the magnified estimates among Moor women may be due to greater exposure to conflict together with experiences of higher socioeconomic deprivation rather than characteristics of ethnicity and should be interpreted with caution.

## Conclusions

While IPV is more likely to occur in the context of socioeconomic disadvantage and post-conflict environments, the complex interplay of poverty, exposure to violence, and more proximal household and clinical factors such as alcohol misuse requires further investigation. Future exploratory studies examining these relationships as well as empirical studies investigating proximal factors is needed, particularly in the absence of an association between SEP and psychological abuse.

## Data Availability

Data is securely stored within the Sri Lankan Department of Census and Statistics. Data is available upon request from the Department.

