## Supplementary Tables for "Socioeconomic and geographic correlates of intimate partner violence in Sri Lanka: Analysis of the 2016 Demographic and Health Survey"

**Supplementary Table 1. Characteristics of respondents versus non-respondents of intimate partner violence questionnaire of Sri Lanka Demographic and Health Survey 2016.**

|  | Respondents N (%) | Non-respondents N (%) | P value^ǂ^ |
| --- | --- | --- | --- |
| *Individual and household factors* |  |  |  |
| Age |  |  |  |
| 15-34 | 6916 (42.2) | 960 (50.3) | <0.001 |
| 35-49 | 9477 (57.8) | 949 (49.7) |  |
| Religion |  |  |  |
| Buddhist | 11731 (71.6) | 1272 (66.7) | <0.001 |
| Hindu | 1856 (11.3) | 222 (11.6) |  |
| Islam | 1529 (9.3) | 243 (12.7) |  |
| Christian/Other | 1277 (7.8) | 172 (9.0) |  |
| Ethnicity |  |  |  |
| Sinhala | 12537 (76.5) | 1391 (72.8) | <0.001 |
| Tamil | 2376 (14.5) | 278 (14.5) |  |
| Moor | 1422 (8.7) | 238 (12.5) |  |
| Other | 58 (0.4) | 3 (0.1) |  |
| Marital status |  |  |  |
| Married | 14979 (91.4) | 1566 (82.1) | <0.001 |
| Living with a man | 662 (4.0) | 50 (2.6) |  |
| Divorced/Separated/Husband died | 753 (4.6) | 293 (15.3) |  |
| Educational attainment |  |  |  |
| Higher education | 780 (4.8) | 76 (4.0) | <0.001 |
| Secondary education | 14283 (87.1) | 1621 (84.9) |  |
| No education/primary | 1330 (8.1) | 211 (11.1) |  |
| Partner’s educational attainment |  |  |  |
| Higher education | 547 (4.0) | 41 (3.1) | 0.03 |
| Secondary education | 11703 (84.6) | 1101 (83.0) |  |
| No education/primary | 1583 (11.4) | 185 (13.9) |  |
| Occupational status |  |  |  |
| Professional | 1204 (7.3) | 124 (6.5) | 0.13 |
| Service | 1550 (9.5) | 192 (10.1) |  |
| Manual labour | 2608 (15.9) | 339 (17.8) |  |
| Unemployed | 11032 (67.3) | 1254 (65.7) |  |
| Partner’s occupational status* |  |  |  |
| Professional | 2429 (18.6) | 164 (13.2) | <0.001 |
| Service | 3611 (27.6) | 405 (32.5) |  |
| Manual labour | 6648 (50.8) | 606 (48.6) |  |
| Unemployed | 396 (3.0) | 71 (5.7) |  |
| Enough income for daily living expenses** |  |  |  |
| Yes or employed | 8484 (76.9) | 912 (72.7) | 0.002 |
| No | 2548 (23.1) | 342 (27.3) |  |
| Household wealth index |  |  |  |
| 1 - Richest | 3214 (19.6) | 348 (18.2) | 0.30 |
| 2 | 3436 (21.0) | 380 (19.9) |  |
| 3 | 3439 (21.0) | 399 (20.9) |  |
| 4 | 3301 (20.1) | 394 (20.7) |  |
| 5 - Poorest | 3003 (18.3) | 387 (20.3) |  |
| *Community-level factors* |  |  |  |
| Sector |  |  |  |
| Urban | 2562 (15.6) | 297 (15.6) | 0.71 |
| Rural | 13204 (80.5) | 1529 (80.1) |  |
| Estate | 627 (3.8) | 82 (4.3) |  |
| Neighbourhood poverty |  |  |  |
| Low | 9103 (55.5) | 1060 (55.5) | 1.00 |
| High | 7290 (44.5) | 849 (44.5) |  |
| Post-conflict residence |  |  |  |
| No | 14113 (86.1) | 1702 (89.2) | <0.001 |
| Yes | 2281 (13.9) | 207 (10.8) |  |

ǂChi-square test of independence was employed to derive p-values.
*Women divorced/separated/husband died were not asked this question.
**Women working in a job or business were not asked this question.

**Supplementary Table 2. Weighted prevalence of intimate partner violence (IPV) in the past year among ever-partnered women aged 15-49 years, by province and district in Sri Lanka**

| Name | Area | Total N | % IPV (95% CI) |
| --- | --- | --- | --- |
| Western | **Province** | **4134** | **12.4 (11.2-13.7)** |
| Colombo | District | 1614 | 14.4 (12.2-16.9) |
| Gampaha | District | 1559 | 12.8 (11.0-14.9) |
| Kalutara | District | 961 | 8.4 (6.7-10.5) |
| Central | **Province** | **2069** | **23.7 (21.4-26.2)** |
| Kandy | District | 1100 | 25.0 (21.8-28.6) |
| Matale | District | 431 | 30.1 (25.9-34.8) |
| Nuwara Eliya | District | 538 | 15.9 (11.8-21.0) |
| Southern | **Province** | **1997** | **16.2 (14.3-18.4)** |
| Galle | District | 803 | 21.4 (18.1-25.2) |
| Hambantota | District | 515 | 5.8 (4.0-8.3) |
| Matara | District | 679 | 17.9 (14.9-21.5) |
| Northern | **Province** | **792** | **34.9 (31.6-38.3)** |
| Jaffna | District | 433 | 39.8 (34.5-45.3) |
| Kilinochchi | District | 87 | 50.4 (45.1-55.7) |
| Mannar | District | 76 | 21.1 (16.3-26.8) |
| Mullaitivu | District | 74 | 29.3 (24.5-34.6) |
| Vavuniya | District | 122 | 18.5 (14.5-23.2) |
| Eastern | **Province** | **1488** | **35.2 (32.0-38.4)** |
| Ampara | District | 666 | 27.1 (23.2-31.4) |
| Batticaloa | District | 490 | 49.9 (44.6-55.1) |
| Trincomalee | District | 332 | 29.5 (24.0-35.7) |
| North-Western | **Province** | **2057** | **12.1 (10.6-13.8)** |
| Kurunegala | District | 1447 | 10.3 (8.7-12.2) |
| Puttalam | District | 610 | 16.5 (13.5-19.9) |
| North-Central | **Province** | **1242** | **8.3 (6.7-10.4)** |
| Anuradhapura | District | 867 | 7.7 (5.7-10.4) |
| Polonnaruwa | District | 375 | 9.8 (7.2-13.0) |
| Uva | **Province** | **1080** | **9.2 (7.5-11.3)** |
| Badulla | District | 649 | 10.4 (8.1-13.1) |
| Moneragala | District | 431 | 7.5 (5.2-10.8) |
| Sabaragamuwa | **Province** | **1534** | **11.9 (10.2-14.0)** |
| Kegalle | District | 538 | 9.0 (6.4-12.7) |
| Ratnapura | District | 996 | 13.5 (11.3-16.1) |
| Total | **National** | **16,393** | **16.9 (16.1-17.6)** |

CI=Confidence Interval.

**Supplementary Table 3. Any intimate-partner violence in the past year among ever-partnered women aged 15-49 years, Sri Lanka: adjusted associations with socioeconomic factors and ethnicity, stratified by post-conflict residence.**

|  | Post-conflict residence | | | Other residence | | |  |
| --- | --- | --- | --- | --- | --- | --- | --- |
|  | **IPV**  **N (%)** | **No IPV**  **N (%)** | **OR (95% CI)** | **IPV**  **N (%)** | **No IPV**  **N (%)** | **OR (95% CI)** | **P value for interaction** |
| Educational attainment |  |  |  |  |  |  |  |
| Higher education | 27 (3.4) | 80 (5.4) | 1.00 | 72 (3.6) | 601 (4.9) | 1.00 | 0.47 |
| Secondary education | 616 (77) | 1230 (83.1) | 1.68 (1.05-2.67) | 1652 (84.2) | 10785 (88.8) | 1.31 (0.97-1.76) |  |
| No schooling/primary education | 157 (19.6) | 171 (11.6) | 3.00 (1.79-5.01) | 239 (12.2) | 764 (6.3) | 2.54 (1.79-3.60) |  |
| Partner's educational attainment |  |  |  |  |  |  |  |
| Higher education | 24 (3.6) | 68 (5.8) | 1.00 | 43 (2.6) | 413 (4) | 1.00 | 0.84 |
| Secondary education | 458 (70.9) | 945 (81.1) | 1.69 (1.03-2.75) | 1305 (79.2) | 8995 (86.7) | 1.42 (0.97-2.07) |  |
| No schooling/primary education | 165 (25.5) | 152 (13.1) | 3.65 (2.13-6.28) | 301 (18.3) | 965 (9.3) | 2.98 (1.99-4.48) |  |
| Occupational status |  |  |  |  |  |  |  |
| Professional | 15 (1.9) | 52 (3.5) | 1.00 | 158 (8.1) | 978 (8) | 1.00 | 0.003 |
| Service | 51 (6.4) | 107 (7.2) | 1.52 (0.89-2.60) | 198 (10.1) | 1194 (9.8) | 1.06 (0.81-1.38) |  |
| Manual labour | 117 (14.6) | 188 (12.7) | 2.14 (1.24-3.70) | 413 (21.1) | 1890 (15.6) | 1.32 (1.04-1.68) |  |
| Unemployed | 617 (77.1) | 1135 (76.6) | 1.95 (1.17-3.27) | 1193 (60.8) | 8088 (66.6) | 0.95 (0.77-1.16) |  |
| Partner's occupational status* |  |  |  |  |  |  |  |
| Professional | 76 (12) | 125 (11.3) | 1.00 | 273 (17.6) | 1955 (20) | 1.00 | 0.009 |
| Service | 75 (11.9) | 191 (17.3) | 0.67 (0.45-1.00) | 444 (28.5) | 2901 (29.6) | 1.07 (0.88-1.30) |  |
| Manual labour | 467 (73.4) | 754 (68.1) | 1.11 (0.79-1.55) | 774 (49.7) | 4653 (47.6) | 1.14 (0.95-1.36) |  |
| Unemployed | 17 (2.7) | 37 (3.4) | 1.05 (0.53-2.05) | 65 (4.2) | 276 (2.8) | 1.57 (1.07-2.30) |  |
| Enough income for daily living expenses** |  |  |  |  |  |  |  |
| Yes | 440 (71.4) | 959 (84.5) | 1.00 | 737 (61.8) | 6348 (78.5) | 1.00 | 0.64 |
| No | 176 (28.6) | 176 (15.5) | 2.66 (2.09-3.38) | 456 (38.2) | 1740 (21.5) | 2.36 (2.03-2.74) |  |
| Household wealth index |  |  |  |  |  |  |  |
| 1 - Richest | 40 (5) | 136 (9.2) | 1.00 | 345 (17.6) | 2694 (22.2) | 1.00 | 0.87 |
| 2 | 82 (10.3) | 257 (17.4) | 1.08 (0.69-1.69) | 361 (18.4) | 2736 (22.5) | 1.07 (0.89-1.29) |  |
| 3 | 103 (12.9) | 269 (18.2) | 1.28 (0.81-2.00) | 376 (19.2) | 2690 (22.1) | 1.17 (0.97-1.42) |  |
| 4 | 179 (22.4) | 335 (22.6) | 1.66 (1.10-2.50) | 416 (21.2) | 2371 (19.5) | 1.52 (1.25-1.84) |  |
| 5 - Poorest | 394 (49.3) | 484 (32.7) | 2.33 (1.60-3.40) | 465 (23.7) | 1659 (13.7) | 2.40 (1.98-2.91) |  |
| Community-level poverty |  |  |  |  |  |  |  |
| Low | 165 (20.7) | 466 (31.5) | 1.00 | 1120 (57.1) | 7351 (60.5) | 1.00 | 0.04 |
| High | 634 (79.3) | 1015 (68.5) | 1.38 (1.09-1.75) | 842 (42.9) | 4799 (39.5) | 1.12 (0.98-1.29) |  |
| Ethnicity*** |  |  |  |  |  |  |  |
| Sinhala | 54 (6.7) | 332 (22.5) | 1.00 | 1655 (84.5) | 10497 (86.7) | 1.00 | <0.001 |
| Tamil | 573 (71.9) | 845 (57.2) | 1.39 (0.10-19.50) | 194 (9.9) | 765 (6.3) | 1.63 (1.06-2.50) |  |
| Moor | 170 (21.3) | 301 (20.4) | 3.62 (0.24-53.47) | 110 (5.6) | 840 (6.9) | 0.78 (0.27-2.25) |  |

OR = Odds Ratio adjusted for age and religion; CI = Confidence Interval.
*Women divorced/separated/husband died were not asked this question.
**Women working in a job or business were not asked this question.
***Ethnicity excludes ‘other’ minority ethnic groups (N=58).

**Supplementary Table 4. Any intimate-partner violence in the past year among ever-partnered women aged 15-49 years, Sri Lanka: sensitivity analysis using complete data (N=13,837).**

|  | IPV (N=2388)  N (%) | No IPV (N=11,450)  N (%) | OR (95% CI) |
| --- | --- | --- | --- |
| *Individual level factors* |  |  |  |
| Age |  |  |  |
| 15-34 | 920 (38.5) | 4691 (41) | 1.00 |
| 35-49 | 1467 (61.5) | 6759 (59) | 1.22 (1.11-1.37) |
| Ethnicity |  |  |  |
| Sinhala | 1453 (60.9) | 9067 (79.2) | 1.00 |
| Tamil | 681 (28.5) | 1386 (12.1) | 3.16 (2.28-4.40) |
| Moor | 248 (10.4) | 952 (8.3) | 3.58 (1.96-6.54) |
| Other | 6 (0.2) | 44 (0.4) | 1.23 (0.42-3.60) |
| Religion |  |  |  |
| Buddhist | 1364 (57.1) | 8451 (73.8) | 1.00 |
| Hindu | 540 (22.6) | 1088 (9.5) | 3.09 (2.65-3.61) |
| Islam | 259 (10.8) | 1024 (8.9) | 1.52 (1.23-1.88) |
| Christian/Other | 225 (9.4) | 886 (7.7) | 1.52 (1.25-1.86) |
| Marital status |  |  |  |
| Married | 2117 (88.7) | 10383 (90.7) | 1.00 |
| Living with a man | 75 (3.2) | 510 (4.5) | 0.81 (0.60-1.11) |
| Divorced/Separated/Husband died | 195 (8.2) | 557 (4.9) | 1.56 (1.27-1.92) |
| Educational attainment |  |  |  |
| Higher education | 85 (3.6) | 589 (5.1) | 1.00 |
| Secondary education | 1952 (81.8) | 10011 (87.4) | 1.37 (1.04-1.80) |
| No education/primary | 350 (14.7) | 850 (7.4) | 2.39 (1.74-3.27) |
| Partner’s educational attainment |  |  |  |
| Higher education | 66 (3) | 475 (4.4) | 1.00 |
| Secondary education | 1665 (76) | 9318 (85.6) | 1.37 (1.01-1.87) |
| No education/primary | 461 (21) | 1098 (10.1) | 2.88 (2.06-4.03) |
| Occupational status |  |  |  |
| Professional | 145 (6.1) | 898 (7.8) | 1.00 |
| Service | 221 (9.3) | 1105 (9.6) | 1.22 (0.94-1.60) |
| Manual labour | 473 (19.8) | 1822 (15.9) | 1.49 (1.16-1.90) |
| Unemployed | 1548 (64.8) | 7625 (66.6) | 1.19 (0.95-1.48) |
| Partner’s occupational status |  |  |  |
| Professional | 350 (15.9) | 2080 (19.1) | 1.00 |
| Service | 519 (23.7) | 3092 (28.4) | 0.97 (0.81-1.16) |
| Manual labour | 1241 (56.6) | 5407 (49.6) | 1.20 (1.03-1.41) |
| Unemployed | 83 (3.8) | 314 (2.9) | 1.47 (1.05-2.07) |
| Enough income for daily living expenses |  |  |  |
| Yes | 991 (64.0) | 5907 (77.5) | 1.00 |
| No | 557 (36.0) | 1719 (22.5) | 2.17 (1.89-2.50) |
| *Household level factors* |  |  |  |
| Household wealth index |  |  |  |
| 1 - Richest | 313 (13.1) | 2398 (20.9) | 1.00 |
| 2 | 385 (16.1) | 2441 (21.3) | 1.23 (1.01-1.48) |
| 3 | 409 (17.1) | 2425 (21.2) | 1.34 (1.10-1.63) |
| 4 | 514 (21.5) | 2287 (20.0) | 1.77 (1.47-2.15) |
| 5 - Poorest | 767 (32.1) | 1898 (16.6) | 2.73 (2.26-3.29) |
| *Geographical and community level factors* |  |  |  |
| Sector |  |  |  |
| Urban | 444 (18.6) | 1798 (15.7) | 1.00 |
| Rural | 1847 (77.4) | 9212 (80.5) | 0.93 (0.79-1.11) |
| Estate | 96 (4.0) | 439 (3.8) | 0.44 (0.31-0.62) |
| Province |  |  |  |
| Western | 460 (19.3) | 3223 (28.1) | 1.00 |
| Central | 420 (17.6) | 1254 (10.9) | 2.38 (1.93-2.95) |
| Southern | 271 (11.4) | 1401 (12.2) | 1.45 (1.17-1.82) |
| Northern | 260 (10.9) | 466 (4.1) | 3.09 (2.39-4.01) |
| Eastern | 445 (18.6) | 751 (6.6) | 4.21 (3.35-5.30) |
| North-Western | 219 (9.2) | 1539 (13.4) | 1.01 (0.81-1.25) |
| North-Central | 59 (2.5) | 849 (7.4) | 0.51 (0.36-0.72) |
| Uva | 86 (3.6) | 790 (6.9) | 0.76 (0.57-1.00) |
| Sabaragamuwa | 168 (7.0) | 1177 (10.3) | 1.01 (0.79-1.28) |
| Neighbourhood poverty |  |  |  |
| Low | 1102 (46.2) | 6591 (57.6) | 1.00 |
| High | 1285 (53.8) | 4858 (42.4) | 1.34 (1.18-1.53) |
| Post-conflict residence |  |  |  |
| No | 1682 (70.4) | 10233 (89.4) | 1.00 |
| Yes | 706 (29.6) | 1217 (10.6) | 3.14 (2.64-3.74) |

OR = Odds Ratio adjusted for age and religion; CI = Confidence Interval.
*Women divorced/separated/husband died were not asked this question.
**Women working in a job or business were not asked this question.
